# The I-KID Study Protocol: Evaluation of Efficacy, Outcomes and Safety of a New Infant Haemodialysis and Ultrafiltration Machine in Clinical Use: a Randomised Clinical Investigation using a Cluster Stepped-wedge Design

**DOI:** 10.1101/2021.07.13.21260443

**Authors:** Heather J Lambert, Shriya Sharma, John NS Matthews, on behalf of the IKID Protocol Development Group

**Affiliations:** The Great North Children’s Hospital, Newcastle Hospitals NHS Trust; Newcastle Clinical Trials Unit, Newcastle University; Department of Mathematics, Statistics & Physics and Population Health Sciences Institute, Newcastle University

**Keywords:** Infant, Kidney, Dialysis, Filtration

## Abstract

**Introduction:** The I-KID study aims to determine the clinical efficacy, outcomes and safety of a novel non-CE-marked infant haemodialysis machine, the Newcastle Infant Dialysis Ultrafiltration System (NIDUS), compared to currently available therapy in the UK. NIDUS is specifically designed for renal replacement therapy in small babies between 0.8 and 8 kilograms.

**Methods and analysis:** The clinical investigation is taking place in six UK centres. This is a randomised clinical investigation using a cluster stepped-wedge design. The study aims to recruit 95 babies requiring renal replacement therapy in paediatric intensive care units over 20 months.

**Registration:** IRAS ID number: 170481

MHRA Reference: CI/2017/0066

ISRCT Number: 13787486

CPMS ID number: 36558

NHS REC reference: 16/NE/0008

Eudamed number: CIV-GB-18-02-023105

Link to full protocol v6.0: https://fundingawards.nihr.ac.uk/award/14/23/26

**What is known about this subject:** Babies in paediatric intensive care unit (PICU) may develop acute renal failure and require therapeutic support.

Renal replacement options for babies are limited because of their small size and limitations of technology.

Current haemodialysis and filtration systems in use in UK are not recommended or licensed for children under 8kg.

**What this study hopes to add:** Renal replacement methods for infants under 8kg in PICU will be compared.

The efficacy, outcomes and safety of a new infant haemodialysis device will be assessed.

Usability of the new device in normal clinical settings outside of the development centre will be examined.

## Introduction

Young babies requiring renal replacement therapy (RRT) present specific therapeutic challenges because of their small size and the current technology available. Publications indicate similar problems faced by clinicians worldwide who use adult devices because of lack of alternatives, and the need for solutions including improved device technology.

This clinical investigation protocol is designed to determine the clinical efficacy, outcomes and safety of a novel non-CE-marked infant haemodialysis machine, the Newcastle Infant Dialysis Ultrafiltration System (NIDUS), compared to currently available RRT in the UK. NIDUS is specifically designed for use in babies between 0.8 and 8 kilograms (kg).There is evidence from a previous single centre pilot study to anticipate NIDUS has the potential to contribute significant benefits to the health of small babies needing RRT [1].

The proposed clinical investigation is a result of a multicentre collaboration between clinicians, scientists, academics, with significant parent and public involvement, throughout its development; working with a manufacturing company, Allmed. The results will have potential to change clinical practice.

The NIDUS machine uses a smaller circuit volume than current devices. Pilot data from the development centre has suggested management of fluid overload and renal failure is possible for small infants, with the potential for reduced exposure to blood products, and more precise control of ultrafiltration and dialysis [1]. Nurses have reported ease of use of the NIDUS within the design centre but this requires evaluation in standard clinical environments.

## Background

There are several populations of babies requiring RRT. Those included in this study are unwell infants in paediatric intensive care units (PICU), who mostly do not have intrinsic renal disease and therefore have good potential for renal recovery. Many are post-operative, especially post cardiac surgery, whose major problem is an acute kidney insult, fluid overload and poor urine output, and others who are septic or have renal failure as part of multi-organ failure. Although mortality and morbidity in PICU varies and is related to the underlying diagnosis, survival of babies in PICU is worse in those with fluid overload [2] or needing RRT [3], of whom up to 20-40% may die [3,4,5,6,7]. RRT is supportive until kidney recovery and although most survivors are independent of RRT at discharge from PICU, data on chronic renal sequelae are lacking. Children requiring RRT in PICU have been reported to have longer length of stay and required more days of ventilator support [6]. There are over 200 infants per year in the UK receiving treatment with continuous RRT in PICU [8a 8b].

Some babies will be excluded – for example, those with an inborn error of metabolism such as urea cycle defects causing hyperammonaemia, as they require emergency, very rapid removal of toxic metabolites by higher than normal dialysis clearances [9], and babies with severe intrinsic renal disease, which is often congenital, who are usually treated with chronic peritoneal dialysis (PD) at home.

### Current Renal Replacement Therapies

PD is used frequently to support infants after open-heart surgery [3,10]. PD is technically simpler than haemodialysis (HD); there is no lower size limit but complications are common in the smallest patients [2]. Ultrafiltration (UF) is unpredictable, and chemical clearance less efficient, especially in unstable babies who develop splanchnic vasoconstriction and who also risk developing necrotising enterocolitis. This renders PD impossible, as does abdominal surgery and congenital abdominal wall defects. Larger critically ill infants with multi-organ failure are often treated with a variety of continuously delivered haemodialysis (HD) modalities (continuous renal replacement therapy, CRRT) [2,3,7]. Vascular access for HD modalities including Continuous Veno-Venous Haemofiltration (CVVH) is problematic as the size of central venous line (CVL) required for adequate blood flow is disproportionately large for the size of the baby especially when a double lumen line is needed.

Whilst there are no randomised controlled trials in infants, publications indicate recurring themes of difficulties with vascular access and blood flows, fluid balance, rapid clotting, loss of circuits and hypotensive episodes at initiation [4,5,6,7].

Conventional HD and CRRT machines in the UK are used in PICU unlicensed as they are CE marked for use in adults and bigger children. Manufacturers quote fluid balance control as ±30 ml/hour [11], and they therefore are not licensed for babies weighing <8 kg (or approved for use in children of <20 kg in the US). The recommended minimum 7-French, dual-lumen vascular access lines and continuous 40 ml/minute blood flows are difficult to achieve in the smallest babies. Their relatively large circuit volume (50-70ml) produces sudden dilution of blood on commencing treatment if crystalloid primed, and increases the risk of anaemia with circuit loss. Hypotension on connection is a reported problem [4,12,13]. Blood priming risks exposing the baby abruptly to aberrant chemical and pH changes, which are reduced by pre-dialysing the circuit [14]. Exposure to blood transfusions increases the risk of developing tissue-type sensitisation which may be important if renal function does not recover and renal or other solid organ transplant is considered in the future.

There is one CE marked new device for smaller children, the CARPEDIEM, which is not yet available in the UK to enable comparisons [15,16], others, notably in USA, have adapted other adult devices like Aquadex [7,17].

### NIDUS Technology

In 1995, a group in Newcastle designed a novel HD circuit, which operated by different principles i.e. by syringes, and uncoupled the baby’s blood flow capacity from the requirements of the dialysis filter [18]. In 2005, they reported the results of automating this as a miniaturised machine (circuit volume less than 10 ml), with which four babies weighing under 4 kg were treated, using a single-lumen access line, and without the need for blood-priming [19]. This device was subsequently developed into NIDUS [1] which is used as the intervention device in the I-KID study. This clinical investigation will contribute to the current knowledge base and further understanding of the effects of RRT and address the need for improved technology to provide RRT effectively and safely for small babies [20,21].

Safety monitoring is an important focus of this study. The NIDUS makes a downloadable constant recording of all activity data including volumes, flows, pressures, alarms and response to alarms so any alarm or event, however small, can be subsequently analysed. The NIDUS potentially provides a safer way of performing RRT on babies by using a novel circuit that allows precise ultrafiltrate control thus reducing the potential for errors in ultrafiltrate removal that would be trivial for larger children but are not for a baby. Its small circuit volume (<10 ml) potentially avoids the need for blood priming with stored blood which has associated immediate risks and long-term risks of developing sensitising antibodies.

## Methods and analysis

The study aims to evaluate the efficacy and precision of NIDUS in ultrafiltration fluid removal and monitor adverse effects of RRT including use of blood product transfusion (Table1). It will also generate a safety profile in the application of NIDUS in the clinical environment.

### Study Design

The study uses a cluster-randomised standard stepped-wedge (SW) design [22] with four periods and three sequences (Figure 1). The control periods use conventional therapy (PD or CVVH), with NIDUS used in intervention periods. Each site will be trained in setting up and using the NIDUS before switching to an intervention period. The design means that all participating centres will have the chance to use both treatments during the course of the study. PICU nurses will need to be competency assessed before each site can begin using the intervention; 24h on call nurse/clinician will be provided from Newcastle for telephone support.

**Figure 1:**
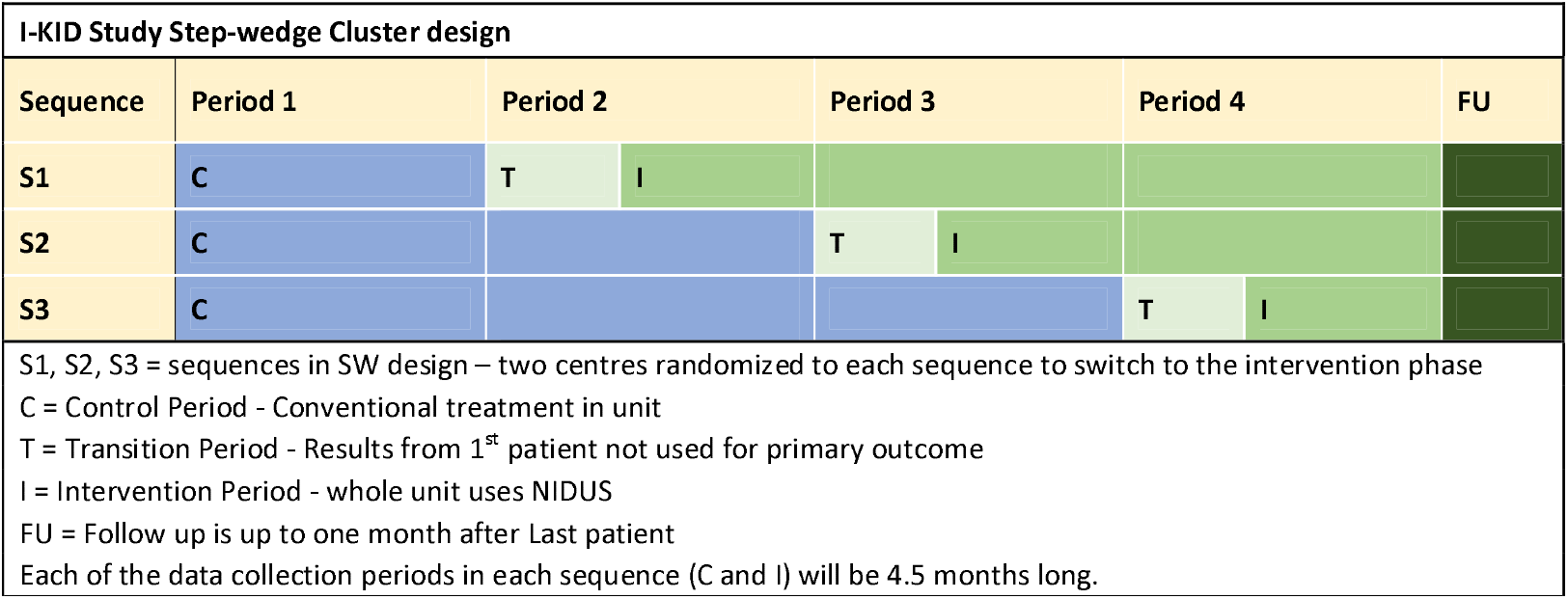
I-KID Study design. I-KID study design sequence (The diagram is indicative. The 4 data collection periods are each 4.5 months. The transition period is up to 2 months and the follow up period for final recruits is one month).

**Figure 2:**
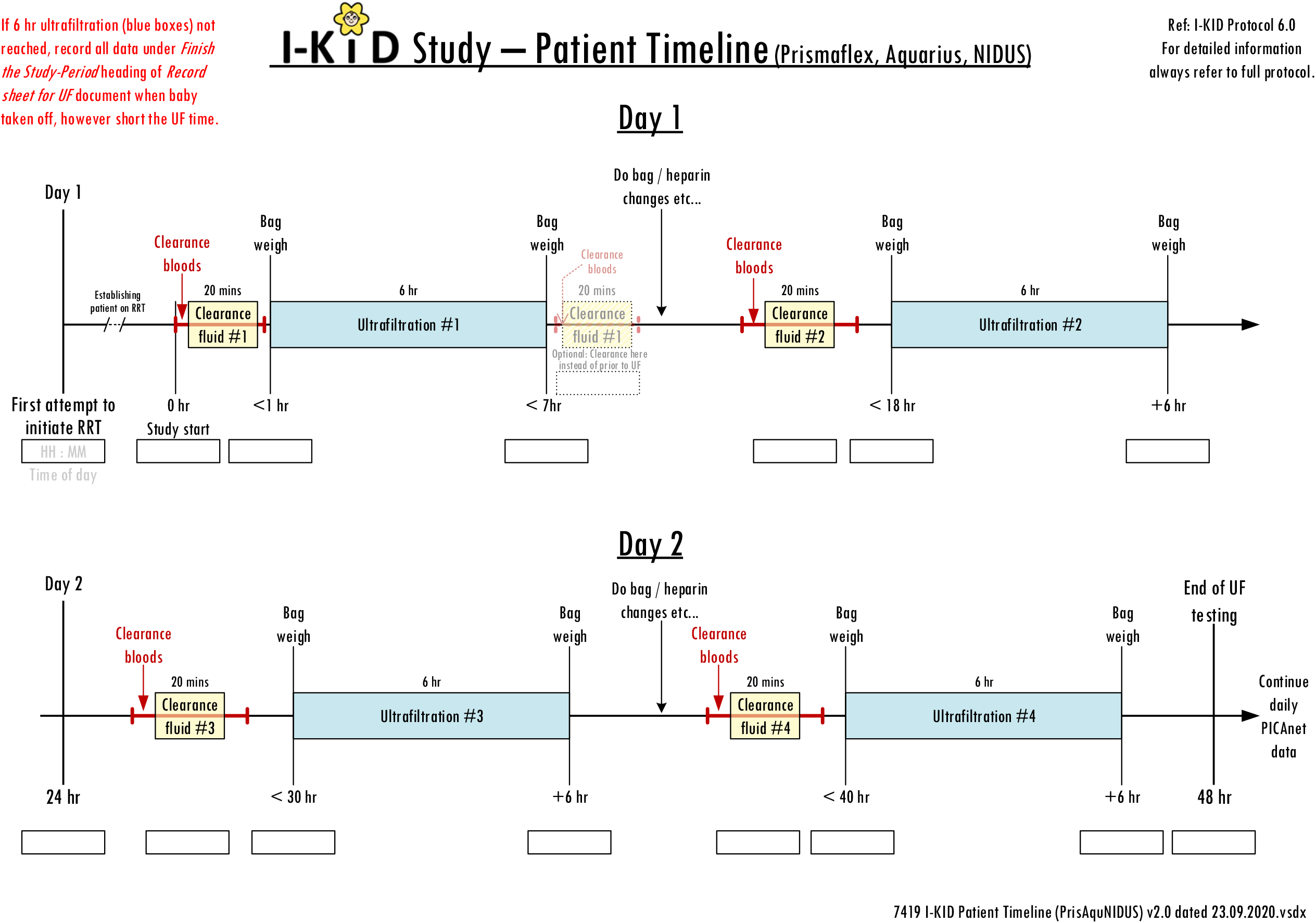
Patient Data Collection Timeline. 2a: Hamodialysis/filtration devices

**2b:**
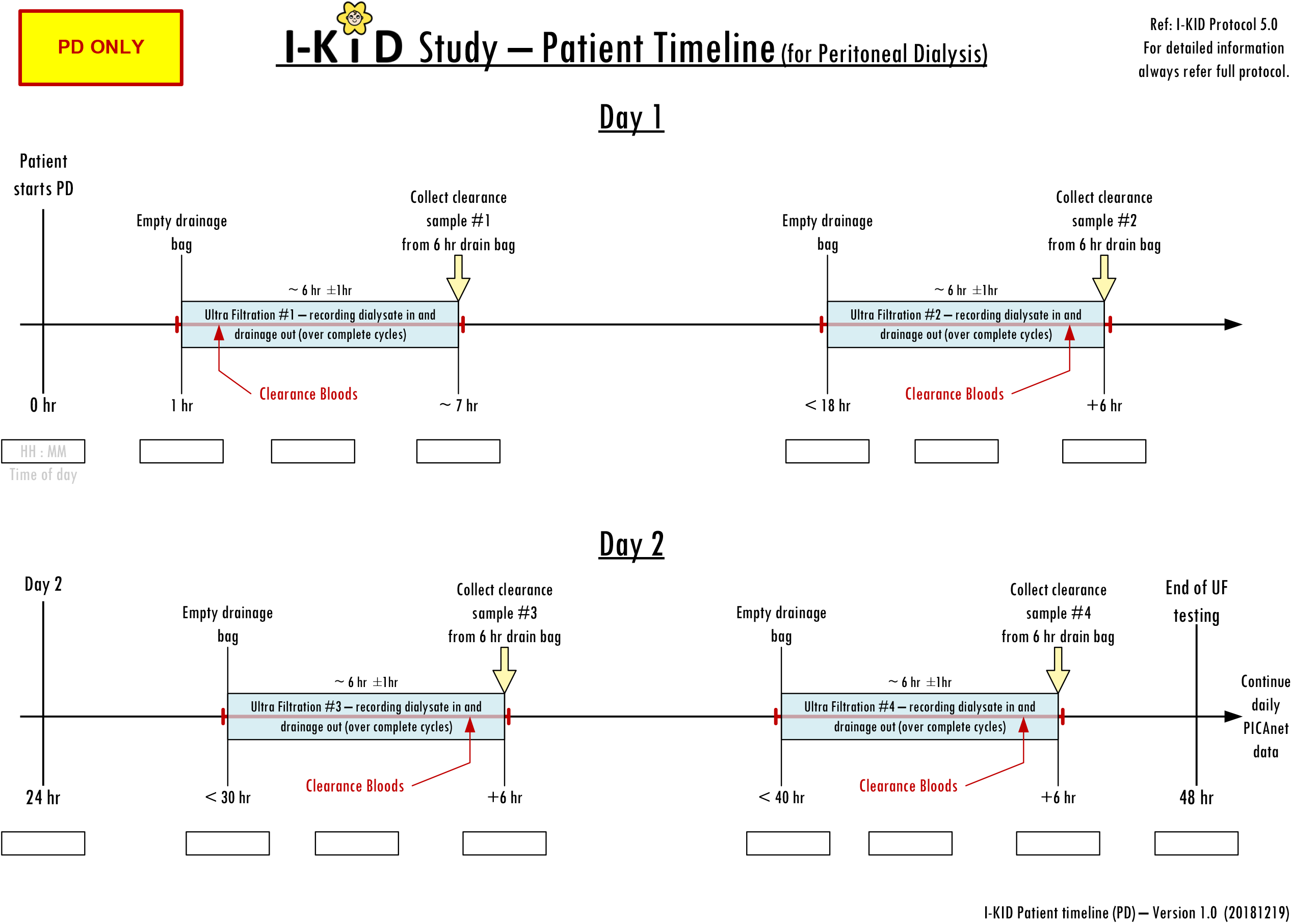
2b: Peritoneal Dialysis

Using a SW design permits the phased training on NIDUS and allows within-centre comparisons to contribute to the treatment estimate.

### Randomisation

Past records suggested that GOSH, Evelina and Southampton (the large centres) treat substantially more patients for RRT than Birmingham, Bristol & Newcastle (the small centres). To avoid large imbalances between the sequences, random permutation in R software was used to allocate one large and one small centre to each sequence. The statistician was blind to the identities of the centres during the allocation.

### Sample selection and outcomes

The study (summarised in Tables 1 and 2) will be conducted in six PICUs. Inclusion and exclusion criteria are shown in Table 3. RRT use and events such as access line changes and blood transfusions will be recorded via the established daily PICANet enhanced renal audit reporting system [8,26]. The weight of the dialysate bags will be measured pre and post dialysis to enable accuracy of fluid removal to be calculated and clearances calculated from measurement of blood and dialysate fluid urea, phosphate and enzymatic creatinine (Figure 3). No additional samples will be taken from the patient for the purposes of this study – only results from routine tests and waste dialysate are needed.

**Table 1:** **I-KID study summary**

|  |  |
| --- | --- |
| **Design** | A multi-centre, randomised clinical investigation using a cluster stepped-wedge design |
| **Study interventions** | Control: current renal replacement therapy (either Peritoneal Dialysis or Continuous Veno-Venous Haemofiltration) |
|  | Experimental intervention: renal replacement therapy using the Newcastle Infant Dialysis Ultrafiltration System (NIDUS) |
| **Objectives** | Primary: To compare the use of a novel haemodialysis device with conventional renal replacement therapy in babies under 8kg treated in Paediatric Intensive Care Units |
|  | Secondary objective: To compare the use of a novel haemodialysis device with conventional renal replacement therapy using the secondary outcome measures |
| **Outcomes** | Primary: Accuracy of fluid removal by technique and compared with prescription |
|  | Secondary:<br>• Haemodynamic status<br>• Biochemical clearances<br>• Number of ventilator free days<br>• Survival<br>• Completion of intended renal replacement therapy course<br>• Need for additional vascular or dialysis access<br>• Unplanned change in circuits<br>• Exposure to blood transfusion<br>• Bleeding events<br>• Anticoagulant use<br>• Parent/Guardian experience<br>• Staff acceptability and usability of device |
| **Study sites** | • Birmingham Childrens Hospital<br>• Bristol Childrens Hospital<br>• Evelina London Childrens Hospital<br>• Great Ormond Street Hospital<br>• Newcastle (Great North Children's Hospital and Freeman)<br>• University Hospitals Southampton |
| **Participants** | Sample: Children 0.8kg to 7.99 kg in PICU who require RRT for renal insufficiency or fluid overload |
|  | Size: approx 95 |
| **Study duration** | Approx. 30 months (approx. 20 months recruitment) |

**Table 2:**
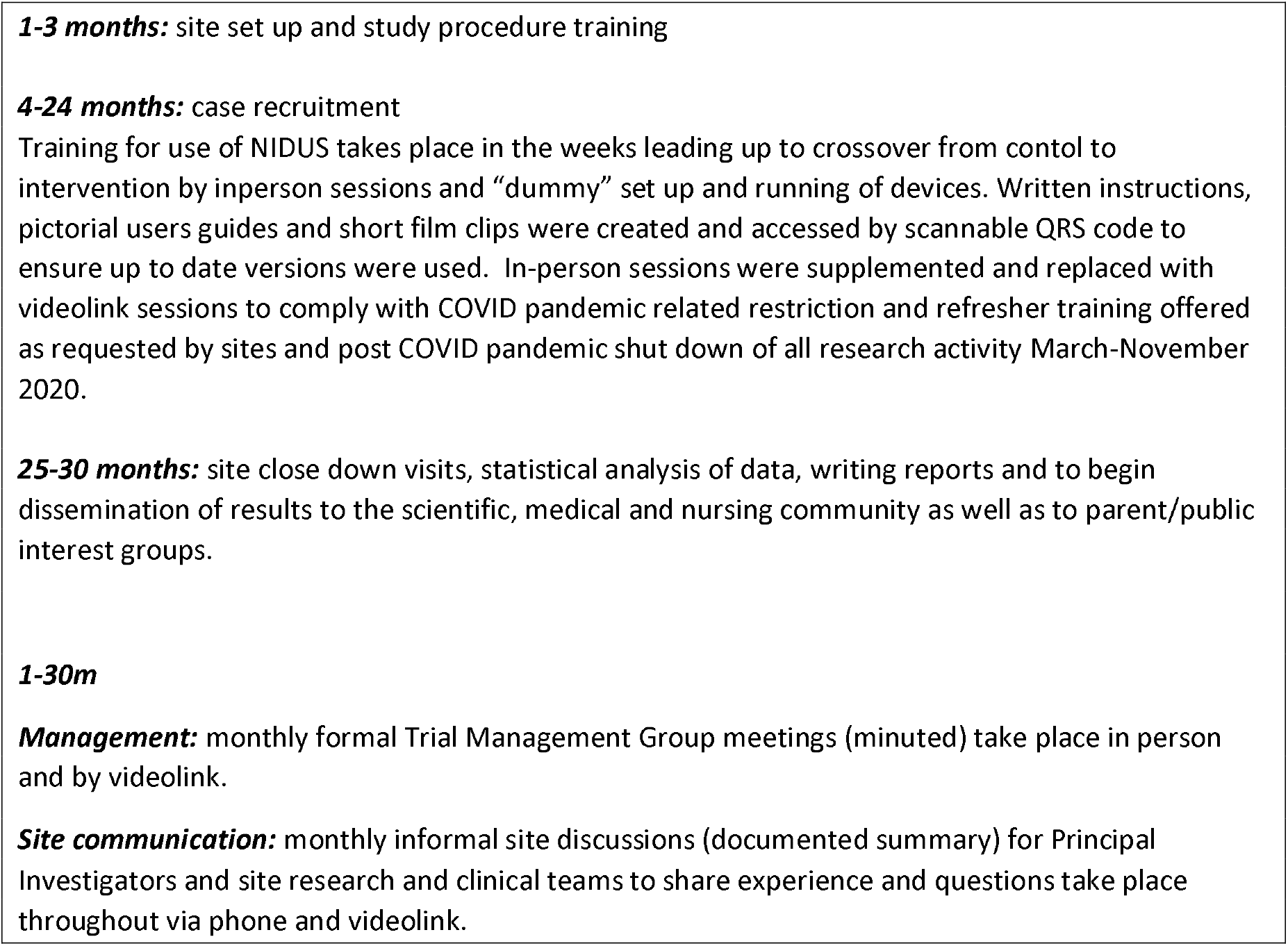
**I-KID Study Timeline**

**Table 3:**
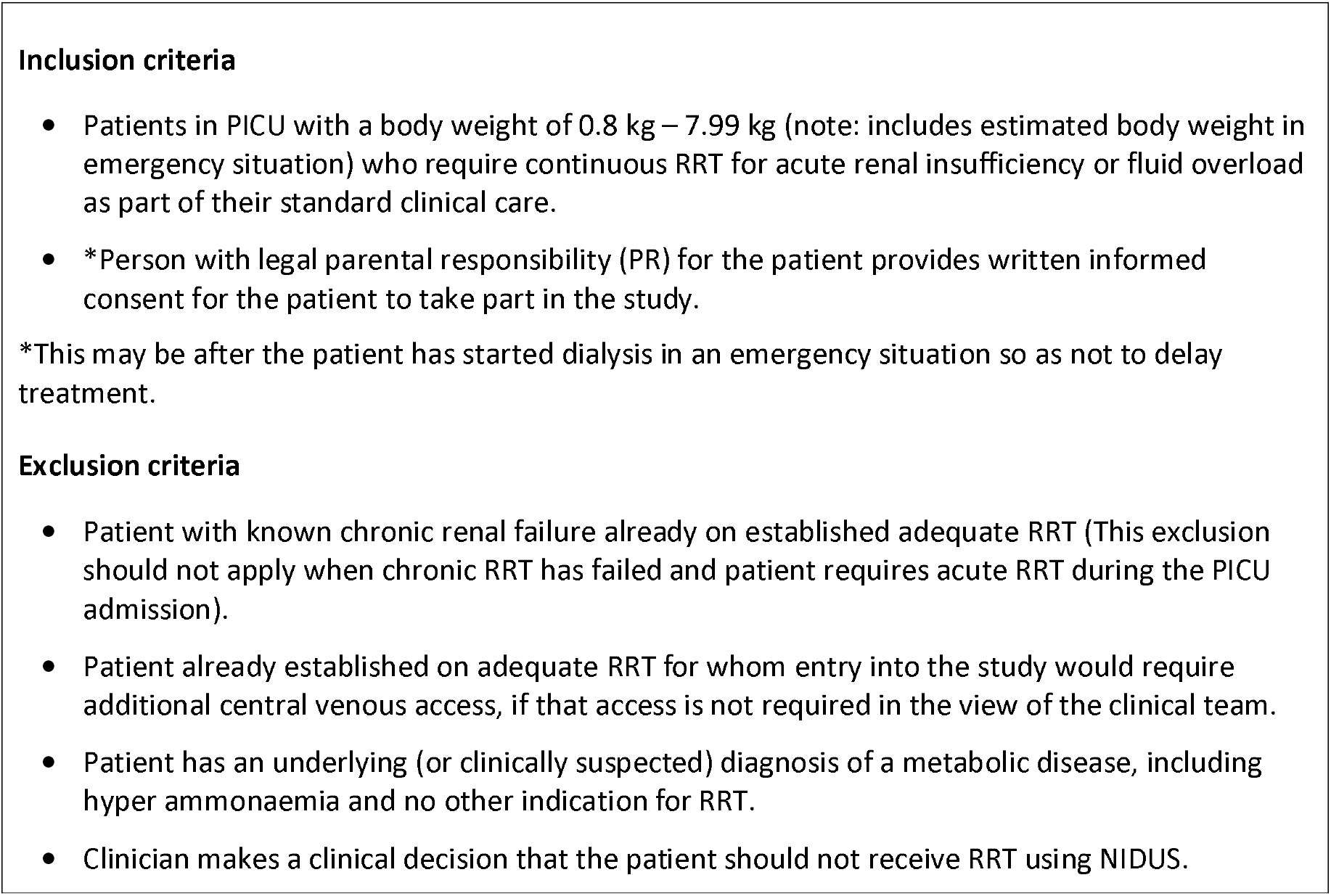
**I-KID Inclusion and Exclusion Criteria**

Using a study-specific questionnaire, parents/guardians will be asked about their experience and staff will be asked about acceptability and usability of the RRT device. Follow up/outcome data will be sought from a routine clinic visit approximately 1 month after start of their RRT; this is to establish whether renal recovery took place: this will include clinical information obtained at discharge from PICU.

### Statistical considerations

#### Primary outcome

The primary aim is to compare the precision of the standard therapy and NIDUS to deliver the fluid removal rate prescribed by the treating physician. The primary outcome is based on the first available determination of fluid removal over a period exceeding one hour and within the first 48 hours of commencement of RRT: if the observed removal is *X* and the prescribed removal is *A*, the primary outcome is log|*X*-*A*|. The expected difference of this quantity between the treatment groups is the log of the ratio of the standard deviations (SDs) of the determinations by the two methods. The method supposes that *X* follows a normal distribution with mean *A* and hence the variance of the outcome is π^2^/8.

#### Sample size

Historical data suggested that annual recruitment to the large centres would be 14 patients, with 9 patients in each of Bristol and Birmingham and 3 in Newcastle. The sample size was determined to detect a ratio of the SDs under the standard therapy and NIDUS of three, with power at least 80% and two-sided type I error of 5%. A three-fold improvement in the precision of fluid removal in this population would be sufficiently marked that it would be likely to change practice. The calculation used the methods in Matthews & Forbes [23], adapted to unequal cluster sizes, and found that four periods in the SW design, each of 4.5 months, gave a power of 80% with an assumed Intraclass Correlation, ICC,of 0.1 and 84% for an ICC of 0.05. It was believed that these represented conservative choices for the ICC.

#### Secondary Outcomes

Fluid removal data aggregated over the duration of RRT, or the first 48 hours if shorter, will be calculated. Biochemical clearances and ventilator-free days while on RRT will be collected. Binary outcomes are: survival [to 30 days and to discharge], haemodynamic status, whether RRT was completed as intended, need for additional vascular access and unplanned change in dialysis circuit, exposure to blood transfusion, bleeding from insertion line and anticoagulant use.

Responses to questionnaires i) parent/guardian about their experience and ii) to staff regarding acceptability and usability.

### Planned Analysis

Analysis of all available data will be on the basis of intention-to-treat. A subgroup analysis will compare NIDUS with conventional CVVH i.e. excluding PD. For the latter group the amount of fluid removed (*X*) will be compared with the amount the machine reports to have been removed *(A)*.

The primary outcome will be analysed using a linear model with fixed effects for treatment, period and cluster. The use of a fixed rather than random effect for cluster is a response to the interruptions to data collection due to the effect of COVID-19 on PICUs and the subsequent difficulty in defining a suitable dispersion structure. Sensitivity analyses will use a generalized estimating equation and will assess the assumptions about *X*. If these are untenable then *X*-*A* will be modelled directly, with treatment dependent variances for the error terms. The above linear model will be applied to the non-binary secondary outcomes. Binary outcomes will be analysed using generalized mixed models if possible but using simple tabulations if more sophisticated analyse are infeasible. Questionnaire data will be tabulated by treatment.

## Ethics and Dissemination

This study is taking place in a high-risk group of sick infants. The design uses cluster-randomisation for reasons of safety, ethics, and acceptability: randomisation by centre, rather than by patient, has been supported by a Research Consumer Group, and in consultation with health professionals and parents. Feedback was sought from a group of parents with children on dialysis in Newcastle where considerable support was given to the study and the design. It was felt that obtaining individual consent for the type of dialysis method to be used would add to families’ stress and anxiety and parents were likely to default to the clinical team for advice. The study design was considered to be a good compromise where the hospital was randomised, with individual consent sought at a later date for collection and recording of information only for the study.

Favourable ethical opinion was obtained from Tyne and Wear South Research Ethics Committee. A letter of no objection was obtained from the Medicines and Healthcare products Regulatory Agency (MHRA).

### Consent

Study Information sheets are provided to parents/guardians of all eligible patients. Tailored consent is obtained appropriate to the phase of the study.

Parent and co-applicant CB has been involved in the study development from the start to ensure that methods are acceptable and sensitive. He took part in multiple teleconference discussions and spoke at the study launch event, and along with other interested parents will take part in dissemination of findings.

A level of urgency to recruit, consent and initiate RRT without compromising the patients’ health further raises ethical concerns [24] and delayed consent will be accepted following CONNECT best practice [25]; consent from bereaved parents may be sought using the bereaved parent/guardian information sheet and consent form. Discussion with the Newcastle Research Consumer Group and individual parents demonstrated how important they felt this study would be. They held favourable views on the study design, use of delayed consent and inclusion of bereaved families (and protocol was amended).

### Safety Reporting

All adverse events (AE), other than those considered consistent with the usual clinical pattern for patients requiring RRT in PICU, and observed Device Deficiency (DD) are collected and recorded. All serious AEs for this study, whether considered device/procedure related or not will be reported to the MHRA in line with regulatory requirements.

### Study Oversight

The study is managed by a Trial Management Group based at Newcastle Clinical Trials Unit (NCTU), with oversight from Study Sponsor, Trial Steering and Data Monitoring Committees. A Safety subcommittee reviews all safety reports.

Data will be handled, computerised and stored in accordance with the Data Protection Act 2018. NCTU will be responsible for the set up and maintenance of the study database and data management procedures.

## Data Availability

Data may be made available once the study is complete.

## Acknowledgements

We thank the parents and families of babies recruited to the study for their important contribution, particularly at a such stressful and anxious time when their baby is unwell.

We acknowledge the work, time and enthusiasm of the site research and clinical staff in making this study happen, their continuous support and for enabling restart to recruitment following interruption due to the COVID pandemic.

We acknowledge the work of Michael Drinnan, Jim Weightman and Alison Bray from the Northern Medical Physics and Clinical Engineering Directorate, NUTH in the technical support and development of the NIDUS device.

## Author’s contributions

- HJL is the chief investigator and conceived the study, led its design and co-ordination and drafted the manuscript.
- MGC conceived the original NIDUS device concept and jointly designed the study and contributed to the writing of the manuscript.
- JNSM jointly designed the study and led the planned statistical analysis and contributed to the writing of the manuscript.
- CB participated in the design of the study and the preparation of parent information and aids in planning and disseminating information about the study.
- DH, QM, JS, KM, CW, RP, LW, JC, RA, EM, DG, JP, AN, HD participated in the design and development of the study and contributed to review of the manuscript.
- MG, SH, NW contributed statistical expertise to protocol development.
- MW, JB, IG & RH contributed to device engineering and study development.
- The NCTU team JWa, SS, E-MH, AS, KR and KN assisted in preparation of the clinical trial protocol to comply with the type recognised by governance and legislative bodies and assisted with the preparation of information sheets and consent forms and overseeing protocol amendments. RW led design of data management.
- DC, JS, RS, VE, JO, CPa contributed to protocol development of training, support and user instruction.
- CPr, KT contributed to protocol development.
- All authors read and approved the final manuscript.

## I-KID Protocol Development Group

Heather Lambert (HL)

John Matthews (JNSM)

Malcolm Coulthard (MGC)

Daljit Hothi (DH)

Quen Mok (QM)

Jon Smith (JS)

Kevin Morris (KM)

Lucy Wirz (LW)

Jean Crosier (JC)

Rachel Agbeko (RA)

Chris Boucher (CB)

Elaine McColl (EM)

David Grant (DG)

John Pappachan (JP)

Andrew Nyman (AN)

Heather Duncan (HD)

Claire Westrope (CW)

PICAnet: Roger Parslow (RP), Richard Feltbower (RW)

Northern Medical Physics & Clinical Engineering Directorate: Michael Whitaker (MW), Joseph Bulmer (JB), Ina Guri (IG), Rebecca Harrison (RH).

Newcastle University Biostatistics Research Group: John Matthews (JM), Michael Grayling (MG), Shaun Hiu (SH), Nina Wilson (NW)

Newcastle Clinical Trials Unit: Jenn Walker (JWa), Shriya Sharma (SS), Ruth Wood (RW), Eva-Maria Holstein (E-MH), Alison Steel (AS), Katherine Rennie (KR) and Karen Nicholson (KN)

NUTH specialist renal nurses: Denise Chisholm (DS), Jayne Straker (JS), Rachel Steel (RS), Vikki Emmet (VE), Julie Office (JO), Christine Pattinson (CPa)

NUTH Sponsor and Project management: Chris Price (CPr), Kelsie Thomas (KT)

## Competing interests

MGC is named on the patent and will receive some royalties if the device goes into commercial production.

The Newcastle upon Tyne NHS Foundation Trust (NUTH) will receive some royalties if the device goes into commercial production.

Allmed are providing 17 NIDUS devices for loan to sites for the I-KID study and for potential compassionate use in the post-study period. NIDUS consumables are purchased as per normal clinical care.

There are no other competing interests declared.

